# Enhancing pain care with the American Pain Society Patient Outcome Questionnaire for use in the Emergency Department (APS-POQ-RED): validating a patient-reported outcome measure

**DOI:** 10.1101/2022.10.17.22280670

**Authors:** James A Hughes, Sarah Hazelwood, Anna-Lisa Lyrstedt, Lee Jones, Nathan J Brown, Rajeev Jarugula, Clint Douglas, Kevin Chu

## Abstract

Pain is the most common symptom experienced when presenting to the emergency department (ED). Estimates indicate over half of all patients will present in pain. EDs typically focus on care process measures, such as time to first analgesic medication. Process-based metrics remove the patient from their own experience. Unfortunately, when patient-reported measures of pain care are used in the ED for quality improvement or research, they vary widely and often lack validation. Previous work has demonstrated that a modified version of the American Pain Society – Patient Outcome Questionnaire – Revised Edition (APS-POQ-R) may provide an ideal patient-reported outcome measure for the adult ED population. However, previous work has left validation incomplete. In this multi-site, multistage research, we demonstrate the construct, convergent and divergent validity and the internal consistency of a modified version of the APS-POQ-R in adult patients presenting to two large, inner-city EDs with moderate to severe acute pain. After three stages of psychometric testing in 646 patients, we present a nine-question, three construct patient-reported outcome measure for moderate to severe pain in the adult emergency department, now known as the American Pain Society – Patient Outcome Questionnaire – Revised for the ED (APS-POQ-RED).

**Perspective:** This article presents the psychometric properties of a revised version of the APS-POQ-R for use in the adult ED. This shortened, ED-specific patient-reported outcome measure (APS-POQ-RED) seeks to provide a standardised, validated measure of patient-reported outcomes of acute pain care in the ED for quality and research purposes.

## Introduction

Pain is the most common symptom in people who present to emergency departments (ED) (1, 2). However, due to a lack of well-validated tools to measure patient-reported outcomes (PROs) of pain care in the ED setting, outcomes of pain care from the perspective of the patient are not well understood (3). This makes it difficult to assess and improve the quality of pain care in the ED. Unidimensional measures of pain intensity are only partially valuable tools because they are limited in scope and fail to account for the multidimensional experience of pain (4-6). Time-based metrics, such as ‘time to be seen’ and ‘time to first analgesic medication’, assume that “faster is better” and provide only a superficial view of the health services’ response to pain rather than the outcomes experienced by the patient (7-9). Furthermore, there are only weak associations between the PROs (such as patient experience) and time-based metrics (9), meaning the patient may receive poor outcomes despite timely care. Previously it has been reported that patients have poor recall of the pain care they receive however have a better recall of the outcomes of this care (10). The symptom of pain and how a patient responds to pain treatment are specific to the individual and best reported by the individual experiencing the symptom (3). Thus, measurement tools are needed to reflect pain’s subjectivity and individual responses to care (11).

In their simple form, a PRO is “a measurement of any aspect of a patient’s health status that comes directly from the patient” (12). When considering a symptom such as pain, PROs offer a measurement of the patient’s perspective of the outcomes of care (13). The incorporation of the patient’s perspective of care can have significant impact on the delivery of effective care and influence change in the manner in which care is delivered (14). While PROs of pain care have been reported in quality and research activities in the ED, the description of their development and validation within this environment is limited. The majority of patient-reported outcomes measures (PROMs) used in the ED are taken directly from other settings without modification or validation (3) which may limit the patients understanding of the instrument and the accuracy of any derived measurements (15).

The most promising and easily modifiable PROM for use in ED is the revised edition of the American Pain Society’s Patient Outcome Questionaire (APS-POQ-R) (3, 16). It encompasses five broad but essential domains of pain: intensity, patient satisfaction, side effects of medications, emotional and physical functioning. Our initial exploratory factor analysis of a modified APS-POQ-R for use in the ED, demonstrated potential construct and discriminate validity based on patient urgency in a single centre (16). The lack of testing of the proposed structure in an additional sample leaves the construct validity and internal consistency incomplete (15). In an effort to overcome these limitations of the previous work (16), and to provide significant evidence of the validity and reliability (15) of a modified APS-POQ-R we conducted a multicentre study of the APS-POQ-R, modified for use in the adult ED using classical test theory.

## Methods

### Aims and Objectives

This study aims to take the previously described adaptation of the APS-POQ-R which has demonstrated incompleate validity, and assesses the construct validity and internal consistancy in adults presenting to the ED with moderate to severe pain. The specific objectives are to:

1. Test the construct validity of a previously described structure (16) of the APS-POQ-R modified for use in adults in the ED.
2. Propose a new construct structure for the modified version of the APS-POQ-R
3. Test the new construct structure proposed in objective two.

### Setting

The study was conducted at the EDs of two principal referral hospitals in Brisbane, Queensland, Australia, with data prospectively collected between September 2021 and January 2022. The Royal Brisbane and Women’s Hospital Emergency and Trauma Centre is an adult ED with 81,761 presentations in 2019 (before the COVID-19 pandemic). The Prince Charles Hospital Emergency Department is a mixed adult and paediatric ED with 100,587 presentations in 2019 (17).

### Instrument

The original APS-POQ-R (18) is an interviewer-administered PROM that was developed for postoperative and cancer-related pain and has been used or adapted to other forms of pain (19), translated into several languages (20-22) and demonstrates cross-cultural validity (23). In studies in the adult ED, the APS-POQ-R has been used as a PROM, either in part (4, 5, 24, 25) or in translated (Danish) entirety (26). However, except for the Danish translation, the validity of the APS-POQ-R in the ED setting has not been fully established. The APS-POQ-R contains two sections, 18 questions that originally mapped to five constructs and a second section that describes the use of non-pharmacological analgesia. Consistent with previously reported validity, this work focuses on the first section (18).

In a previous study (16), we conducted an exploratory factor analysis (EFA) of a version of the APS-POQ-R slightly modified to better suit patients with moderate to severe acute pain in the ED setting, retaining 18 questions from the original APS-POQ-R (18). Those modifications consisted of changes to the wording of questions related to the reporting timeframe, i.e. the phrase “…(in) the first 24 h in the hospital or after your operation” was changed to “…in the emergency department”. There was also an unintentional modification due to a transcription error, in which the question about how much the pain caused the patient to feel anxious was omitted. In the current study, we repeated this EFA by including the question about anxiety. Furthermore, an additional modification was made, in which the two questions relating to the effects of pain on sleep (in the ED) were excluded from the analysis (for objective two and three) because of their lack of relevance to acute pain in the ED setting.

### Participants

Patients were elidgible for inclusion if they presented to ED with acute pain for less than six weeks and had an initial documented or self-reported pain score of 4/10 or greater were eligible. Patients were ineligible if there were triaged into the most urgent category (Australasian Triage Scale, category one), intoxicated with alcohol or other drugs, aged below 18 years, unable to give consent, did not speak English or had cognitive impairment.

### Data collection

For each participant, data were collected by a research nurse directly into an electronic data capture form (RedCap, Vanderbilt University) using a tablet computer following study enrollment. In addition, information that the patient did not provide was collected from the Emergency Department Information System. Data included the participant’s age, sex, mode of arrival to the ED, Charleston comorbidity score, and time from ED arrival to the first analgesic medication.

### Sample and Sample size

We used a convenience sample of patients who met the inclusion criteria. During the shifts of the research nurses (0700 – 1600, Monday to Friday), consecutive ED patients were approached by the research nurse and invited to participate. Patients were only enrolled after their ED care had been completed, either before discharge or while awaiting admission to the hospital. No patients were enrolled once they had left the ED.

The minimum sample size was calculated based on 18 questions over three stages of data analysis. While there is some conjecture around sample size in factor analysis, a sample size of 10 responses per question or an overall sample size of 200 (for less than 40 questions) is deemed adequate (15, 27). Therefore, we aimed to collect 200 participants per objective or 600 participants across the two sites. Funding for research assistants was available for 18 weeks at each site, and while we aimed for a minimum of 200 patients at each site, data collection would continue until the end of the 18 weeks.

### Statistical Analysis

Data were analysed using SPSS (IBM Corp. Released 2021. IBM SPSS Statistics for Windows, Version 28.0. Armonk, NY: IBM Corp). Descriptive statistics were used to summarise the population characteristics. Continuous data were summarised using means and standard deviations. Frequencies and percentages summarised categorical data. The population was split into three even groups using random number allocation (random.org), each group was used to test one of the objectives. ANOVA and the Chi-squared tests were used to determine between-group differences in characteristics (Table 1). Finally, the responses to the 18 questions in the APS-POQ-RED were summarised as means and standard deviations for all patients and each group (Table 2).

**Table 1.** Description of the population and differences between groups.

|  |  | Total | Group One | Group Two | Group Three | Test | P-Value |
| --- | --- | --- | --- | --- | --- | --- | --- |
| Age (mean, SD) |  | 48.3 (19.2) years | 46.9 (19.0) years | 48.8 (19.0) years | 49.35 (19.6) years | F(2,643) = 1.006 | 0.366 |
| Sex (n, %) | | | | | | $\chi^2$ (4) = 3.778 | 0.437 |
|  | Female | 359 (55.6%) | 122 (56.7%) | 121 (56.3%) | 116 (53.7%) |  |  |
|  | Male | 283 (43.8%) | 93 (43.3%) | 93 (43.3%) | 97 (44.9%) |  |  |
|  | Other | 4 (0.6%) | 0 (0.0%) | 1 (0.5%) | 3 (1.4%) |  |  |
| MOA (n, %) | | | | | | $\chi^2$ (4) = 6.249 | 0.181 |
|  | Walk-in | 386 (59.8%) | 131 (60.9%) | 130 (60.5%) | 125 (57.9%) |  |  |
|  | Ambulance Service | 257 (39.8%) | 84 (39.1%) | 85 (39.5%) | 88 (40.7%) |  |  |
|  | Other | 3 (0.5%) | 0 (0.0%) | 0 (0.0%) | 3 (1.4%) |  |  |
| ATS category (n, %) | | | | | | $\chi^2$ (6) = 5.196 | 0.519 |
|  | Two | 97 (15.0%) | 27 (12.6%) | 38 (17.7%) | 32 (14.8%) |  |  |
|  | Three | 369 (57.1%) | 129 (60.0%) | 114 (53.0%) | 126 (58.3%) |  |  |
|  | Four | 165 (25.8%) | 52 (24.2%) | 60 (27.9%) | 53 (24.5%) |  |  |
|  | Five | 15 (2.3%) | 7 (3.3%) | 3 (1.4%) | 5 (2.3%) |  |  |
| Charlson score (mean, SD) |  | 2.0 (2.8) | 1.9 (2.8) | 1.9 (2.6) | 2.2 (2.9) | F(2,643) = 0.839 | 0.432 |
| Receipt of Analgesia (n, %) | | | | | | $\chi^2$ (2) = 1.979 | 0.372 |
|  | Yes | 592 (91.6%) | 197 (91.6%) | 193 (89.8%) | 202 (93.5%) |  |  |
|  | No | 54 (8.4%) | 18 (8.4%) | 22 (10.2%) | 14 (6.5%) |  |  |
| TTA, (mean, SD) minutes |  | 74.1 (73.2) | 73.4 (78.6) | 72.8 (71.9) | 75.9 (69.1) | F(2,589) = 0.097 | 0.907 |

MOA = Mode of arrival, ATS = Australasian Triage Scale, TTA = Time to first analgesic medication, SD = Standard Deviation.

**Table 2:** Summary of responses from the first nine questions of the modified APS-POQ-R.

| Question |  | Total |  | Group One |  |  | Group Two |  |  | Group Three |  |  |
| --- | --- | --- | --- | --- | --- | --- | --- | --- | --- | --- | --- | --- |
|  | N | Mean | SD | N | Mean | SD | N | Mean | SD | N | Mean | SD |
| 1. On this scale, please indicate the least pain that you had in the emergency department. | 646 | 2.26 | 2.32 | 215 | 2.50 | 2.40 | 215 | 2.28 | 2.27 | 216 | 2.01 | 2.29 |
| 2. On this scale, please indicate the worst pain you had in the emergency department. | 646 | 7.78 | 1.90 | 215 | 7.93 | 1.88 | 215 | 7.82 | 1.85 | 216 | 7.60 | 1.95 |
| 3. How often were you in severe pain in the emergency department? Please select the best estimate of the percentage of time you experienced severe pain | 646 | 6.94 | 3.28 | 215 | 3.53 | 3.49 | 215 | 3.00 | 3.13 | 216 | 2.68 | 3.18 |
| 4. Select one number that best describes how much pain interfered or prevented you from: |  |  |  |  |  |  |  |  |  |  |  |  |
| a) Doing activities in bed such as turning, sitting up, repositioning? | 646 | 6.16 | 3.27 | 215 | 6.20 | 3.34 | 215 | 6.23 | 3.27 | 216 | 6.06 | 3.20 |
| b) Doing activities out of bed, such as walking, sitting in a chair, standing at a sink? | 646 | 6.28 | 3.23 | 215 | 6.35 | 3.23 | 215 | 6.32 | 3.19 | 216 | 6.17 | 3.28 |
| c) Falling asleep? | 646 | 4.82 | 3.72 | 215 | 5.29 | 3.73 | 215 | 4.85 | 3.70 | 216 | 4.33 | 3.68 |
| d) Staying asleep? | 646 | 4.37 | 3.79 | 215 | 4.88 | 3.86 | 215 | 4.29 | 3.81 | 216 | 3.96 | 3.67 |
| 5. Pain can affect our mood and emotions. On this scale, please select the one number that best shows how much the pain caused you to feel: |  |  |  |  |  |  |  |  |  |  |  |  |
| a) Depressed? | 646 | 1.48 | 2.87 | 215 | 1.87 | 3.22 | 215 | 1.05 | 2.40 | 216 | 1.52 | 2.89 |
| b) Frightened? | 646 | 2.34 | 3.24 | 215 | 2.35 | 3.27 | 215 | 2.30 | 3.20 | 216 | 2.38 | 3.26 |
| c) Helpless? | 646 | 2.90 | 3.56 | 215 | 2.98 | 3.66 | 215 | 2.95 | 3.55 | 216 | 2.77 | 3.48 |
| d) Anxious | 646 | 3.93 | 3.45 | 215 | 4.06 | 3.52 | 215 | 3.78 | 3.37 | 216 | 3.95 | 3.48 |
| 6. Have you had any of the following side effects? Please select 0 if no. Please circle the number that best shows the severity of each: |  |  |  |  |  |  |  |  |  |  |  |  |
| a) Nausea? | 646 | 2.18 | 2.97 | 215 | 2.22 | 2.99 | 215 | 1.87 | 2.73 | 216 | 2.44 | 3.17 |
| b) Drowsiness? | 646 | 1.00 | 2.22 | 215 | 1.13 | 2.34 | 215 | 0.95 | 2.23 | 216 | 0.91 | 2.08 |
| c) Itching? | 646 | 0.37 | 1.46 | 215 | 0.35 | 1.47 | 215 | 0.39 | 1.41 | 216 | 0.39 | 1.49 |
| d) Dizziness? | 646 | 0.91 | 2.03 | 215 | 1.09 | 2.29 | 215 | 0.75 | 1.77 | 216 | 0.89 | 1.98 |
| 7. In the emergency department, how much relief of your pain did you receive? | 646 | 3.03 | 3.11 | 215 | 3.49 | 3.32 | 215 | 3.01 | 2.99 | 216 | 2.60 | 2.97 |
| 8. Were you allowed to participate in decisions about your pain treatment as much as you wanted to? | 646 | 4.48 | 3.16 | 215 | 4.67 | 3.20 | 215 | 4.38 | 3.15 | 216 | 4.39 | 3.14 |
| 9. Select one number that best shows how satisfied you are with the results of your pain treatment in the emergency department? | 646 | 2.13 | 2.35 | 215 | 2.19 | 2.40 | 215 | 2.28 | 2.36 | 216 | 1.93 | 2.27 |

SD = Standard Deviation. All items were measured on a scale of 0 (0%) to 10 (100%) where 0 equalled a positive outcome (i.e. no pain) to 10 equated to a negative outcome (i.e. worst possible pain). Questions 7,8 and 9 have been recoded to match the direction of the other answers.

The previously described fit of the APS-POQ-R modified for use in the ED was tested with confirmatory factor analysis (CFA) in IBM Amos version 28. The fit of this structure was assessed using the Root Mean Square Error of Approximation (RMSEA), Comparative Fit Index (CFI) and Tucker-Lewis Fit Index (TLI) (28). The model was considered a good fit if the CFI and TLI were greater than 0.9 (29) and the RMSEA was less than 0.08 (30)

EFA (Principal axis factoring) was then conducted using IBM SPSS version 28, with a Promax rotation and Kaiser Normalisation to identify a new structure within the current data. Models with eigenvalues greater than 1, were investigated for the best statistical and clinical fit. Questions were considered loading to a factor if the coefficient was at least 0.4. In addition, the discriminant validity (correlation between factors) and internal consistency (Cronbach alpha) for each factor were reported within the final proposed structure.

The structure of the APS-POQ-R proposed in the EFA was then tested using CFA in IBM Amos version 28. The fit of this structure was assessed using the RMSEA, CFI, and TLI (28) with the previously defined thresholds. Construct validity of the final APS-POQ-R tool was assessed using Composite Reliability (CR) for internal validity (good CR ≥ 0.7), convergent and divergent validity was assessed using Average Variance Extracted (AVE) and Maximum Shared Variance (MSV). Convergent and discriminant validity indicate how individual items correlate with their latent factors. Convergent validity measures the level of variance explained by a construct versus the level due to measurement error and can be judged by AVE≥ 0.5. Whereas discriminant validity is comparing the amount of the variance explained by each construct to the shared variance of other constructs to ensure each construct is measuring different aspects, therefore, MSV should be less than AVE and the square root of AVE greater than inter-construct correlations (31-33).

### Ethical considerations

This study received approval from the Human Research Ethics Committee of the Royal Brisbane and Women’s Hospital (LNR/2019/QRBW/55143). All patients gave written informed consent after explaining the study by one of the research assistants and were free to withdraw from the study until publication.

## Results

A total of 653 patients were recruited for the study across the two sites. One patient withdrew consent after data collection, one was under 18 years at the time of consent, and five who never experienced more than 3/10 pain in the ED were excluded from data analysis. This left a total sample of 646 patients, randomly split into three groups of 215, 215 and 216. Table 1 shows no statistically significant between-group differences in demographic or clinical characteristics. Table 2 summarises the answers to each question of the APS-POQ-R for the three groups and the whole sample.

### Confirmatory Factor Analysis

The previously described structure was tested using CFA on group one as described in the methods. The structure of the model and individual factor loadings can be found in the supplementary material (Supplimentary Figure One). This model did not prove to be a good fit when tested. The CFI reported for this model ranged from 0.605 to 0.715, well outside the 0.900 thresholds considered a good fit. The RMSEA for this model was 0.153 (90% CI 0.143 – 0.164, p<0.001), also well above the 0.08 threshold for a good model fit. Standardised factor loadings ranged from 0.22 to 1.01, with three of the five subscales having factors that loaded less than the 0.7 minimum. Therefore this data had a poor fit for the previously proposed structure. Analysis progressed to identify a new structure using EFA.

### Exploratory factor analysis

An EFA conducted on group two data resulted in a Kaiser-Meyer-Olkin measure of sampling adequacy of 0.792, and a statistically significant Bartlett’s test of sphericity (χ^2^ (120) = 1539.0, p<0.001). A solution with up to five factors (subscales) had an Eigenvalue of greater than 1. Taking into account the structure, questions, and analysis, a three-factor solution, consisting of the pain relief and satisfaction subscale, the affective distress subscale and the pain interferance subscale, and which explained 53.6% of the variation in the data, was selected as the solution to undergo further confirmatory factor analysis. Both the four and five factor solutions split questions into constructs that did not demonstrate clincal applicability or a clear concept. Table 3 shows the internal consistency of each of the three factors, and Table 4 outlines the factors that were omitted. Supplementary Table 1 shows the pattern matrix for the three-factor solution.

**Table 3:** Subscale item to total correlations and Cronbach alpha for a three-factor solution.

|  | Scale<br>Mean if<br>Item<br>Deleted | Scale<br>Variance if<br>Item<br>Deleted | Corrected<br>Item-Total<br>Correlation | Cronbach's<br>Alpha if Item<br>Deleted |
| --- | --- | --- | --- | --- |
| **Pain relief and satisfaction subscale (Cronbach alpha = 0.891)** |  |  |  |  |
| 1. On this scale please indicate the least pain that you had in the emergency department? | 8.29 | 57.12 | 0.776 | 0.861 |
| 3. How often were you in severe pain in the emergency department? | 7.58 | 47.08 | 0.744 | 0.874 |
| 7. In the emergency department, how much relief of your pain did you receive? | 7.56 | 45.31 | 0.860 | 0.821 |
| 9. Select one number that best shows how satisfied you are with the results of your pain treatment in the emergency department. | 8.29 | 57.63 | 0.715 | 0.879 |
| **Affective Distress subscale (Cronbach alpha = 0.823)** |  |  |  |  |
| 5. Pain can affect our mood and emotions. On this scale, please select the one number that best shows how much the pain caused you to feel |  |  |  |  |
| B. Frightened | 6.72 | 39.63 | 0.655 | 0.780 |
| C. Helpless | 6.33 | 35.99 | 0.696 | 0.739 |
| D. Anxious | 5.15 | 36.23 | 0.687 | 0.748 |
| **Pain interference subscale (Cronbach alpha = 0.908)** |  |  |  |  |
| 4. Select one number below that best describes how much pain interfered or prevented you from: |  |  |  |  |
| A. Doing activities in bed such as turning, sitting up, repositioning? | 6.32 | 10.16 | 0.832 | + |
| B. Doing activities out of bed such as walking, sitting in a chair, standing at the sink? | 6.23 | 10.69 | 0.832 | + |

**Table 4:**
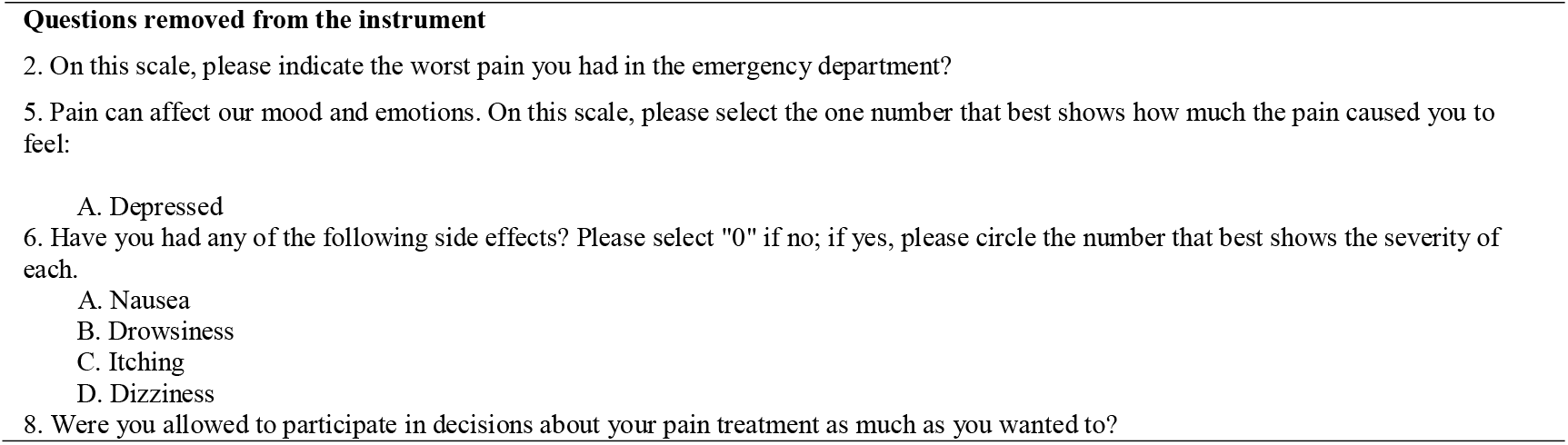
Questions removed from the instrument.

**Table 5:** Validity and Reliability.

| Factor | CR | AVE | MSV |
| --- | --- | --- | --- |
| **Distress** | 0.824 | 0.609 | 0.216 |
| **Pain_Relief** | 0.899 | 0.692 | 0.216 |
| **Pain_Interference** | 0.922 | 0.855 | 0.211 |

CR = Composite reliability, AVE Average Variance Extracted, MSV = Maximum Shared Variance

**Table 6:**
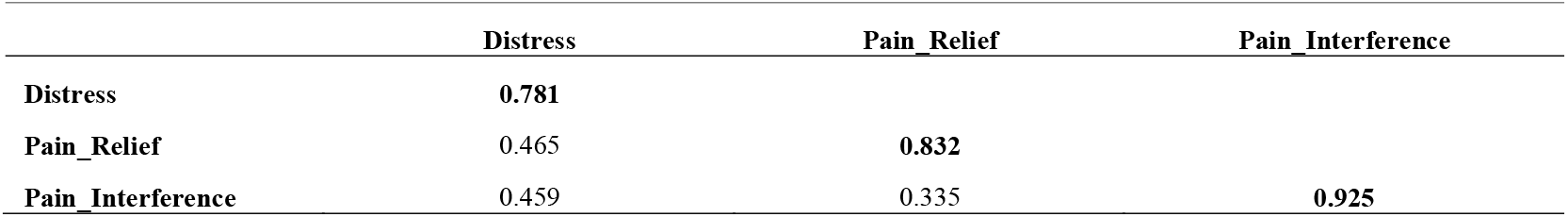
Factor Correlation Matrix with the square root of the average variance explained on the diagional.

|  | **Distress** | **Pain_Relief** | **Pain_Interference** |
| --- | --- | --- | --- |
| **Distress** | **0.781** |  |  |
| **Pain_Relief** | 0.465 | **0.832** |  |
| **Pain_Interference** | 0.459 | 0.335 | **0.925** |

### Confirmatory factor analysis

A CFA was conducted on group three data. The overall structure and coefficients of the analysis are shown in Figure 1. All of the questions loaded to the factors at >0.7. The fit indices indicated that the model was a good fit: TLI = 0.959, and CFI = 0.973, both meeting the minimum threshold of 0.900. The RMSEA was 0.079 (90%CI 0.052 – 0.106, p=0.039) indicating a reasonable fit.

**Figure One:**
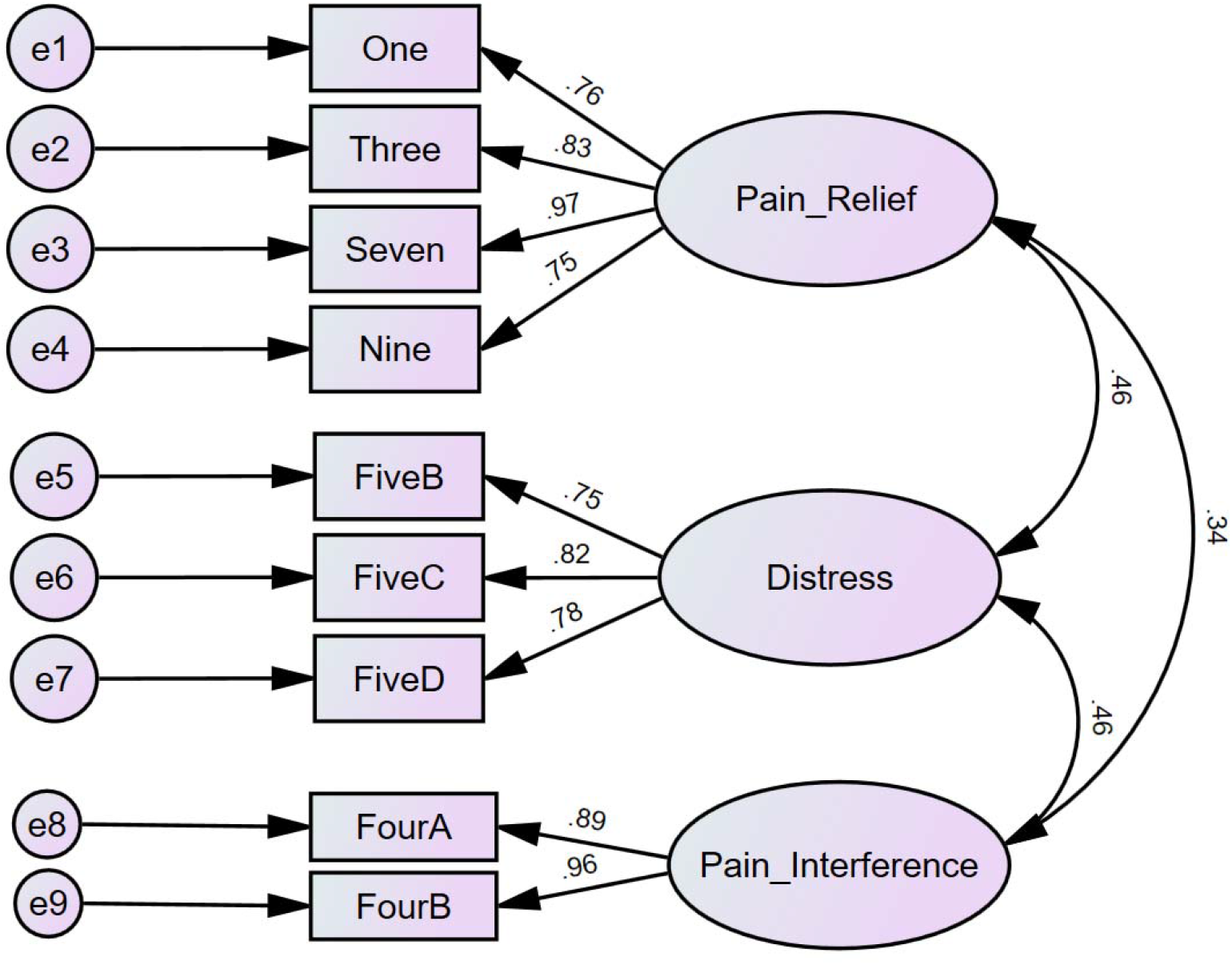
Structure of the three-factor solution showing the factor loading and discriminate ability between constructs. The circles represent the error variance, the rectangles represent the questions included (as per table 2) and the ovals represent the constructs.

Table five and six demonstrate the validity and reliability the model presented. In Table five the composite reliability of the three constructs presented are greater than 0.7 and therefore demonstrate reliability of these constructs (31). The average variance explained for each of the three constructs is greater than 0.5 therefore the three constructs demonstrate convergent validity (33). Discriminant validity is demonstrated in table five where the maximum shared variance is less than the average variance explained and in table six where the square root of the AVE is greater than the inter-construct correlations (32).

## Discussion

In this three-stage prospective validation of the APS-POQ-R for acute pain presenting to the adult ED, we have identified nine questions that map to three latent constructs (subscales). In patients presenting to the ED with moderate to severe acute pain. These ten questions measure the PROs of pain relief and satisfaction, affective distress, and pain interference. This demonstrates the utility of the modified APS-POQ-R in the measurement of PROs of pain care in the adult ED. Our revised edition of the APS-POQ-R for acute pain in the ED will now be known as the APS-POQ-RED (American Pain Society – Patient Outcome Questionnaire – Revised for the ED).

The structure described in the previous adaptation of the APS-POQ-R (16) could not be replicated within this study, despite one of the study locations being the same and replication of the inclusion and exclusion criteria. This highlights the limitations of reporting only EFA as a measure of construct validity without the concurrent use of CFA in a separate sample (15). The generalisability of an EFA model to a CFA model is significantly influenced by several factors, such as the assumption of error within the dataset and the distributional assumption of the data collected (34). The limitations of this earlier work have been overcome in the study presented by ensuring both EFA and CFA are completed on separate samples, collected at the same time, within the same population.

The pain severity and satisfaction subscale is the first of the three latent constructs measured within the revised tool, comprising the least pain severity the patient reported in the ED, how often they were in severe pain, the amount of relief and their overall satisfaction. This construct correlates well with the most commonly measured patient-reported outcomes in the ED literature (pain intensity and patient satisfaction) (16) as well as the two of the constructs measured in the initial development of the APS-POQ-R (pain severity and sleep interference, perceptions of care,) (18). The relief of pain, including the endpoint of pain relief, has been previously shown to be highly correlated with patient satisfaction. Taylor et al., (2015) have shown that targeting adequate analgesia (decrease of at least two points until the pain is rated as less than four on an eleven-point scale) is associated with high levels of patient satisfaction (4, 25). Time to first analgesic medication, as a surrogate measure of the time in severe pain, has been shown to influence patient satisfaction (9). This construct is related to treatment. In future studies of patient-reported pain care outcomes using the APS-POQ-RED in the ED, this construct should be correlated with the treatment given, including time to the first analgesic medication.

The affective distress subscale comprising the degree to which the patient felt frightened, anxious, or helpless because of their pain, was the second subscale identified in the revised tool. Pain rarely exists without co-occurring symptoms such as anxiety (35, 36) Alterations in the affective functioning of the patient can be a result of the pain, the lack of knowledge of the cause or the treatment, and receiving care in an environment that may be unfamiliar and imposing. The relationship between pain and affective distress is well acknowledged within the paediatric ED pain literature, with numerous interventions designed to reduce both (37-42). However, only 19% of PROs reported in the adult literature include emotional functioning as an outcome of pain care (3). A recent review of pain and anxiety measures in the adult ED found that no measure of the co-occurring symptoms of anxiety and pain had been reported in practice outside of quality improvement or research spheres (36). The acknowledgement of affective (emotional) distress as an outcome of effective pain care in the adult ED is the first step in acknowledging co-occurring symptoms (or symptom clusters) within the adult ED.

The third subscale identified within the tool is the pain interference subscale. This subscale identified the impact of pain on the patient’s physical functioning through its two questions. Pain is well known to impact the patient’s ability to function; however, this relationship is not linear as seemingly minor pain can impair a patient’s function (6). Reduction in function can often be the precursor to patients seeking care in an ED. A lack of functional improvement secondary to disease progression is a leading cause of representation (43, 44).

The instrument presented in this work has several uses within the ED. The first use is to measure the outcomes of interventional and quality improvement studies in a consistent manner that is comparable across studies. With an increasing focus on the pharmacological treatment of pain in the ED the use of objective PROMS in interventional studies, especially opiate alternative, and sparing projects, allows the patient voice to compliment other outcomes of the intervention. The second way this instrument may be useful is in identifying the relationship between process/health service outcomes of pain care in the ED and those reported by the patient. Previous work in this area has demonstrated a poor relationship between process measures and patient centred outcomes, however the outcomes measures used were limited and not validated for acute pain in the ED (9).

## Conclusion

In this work we have demonstrated the structural validity and internal consistency of a modified version of the APS-POQ-R for use in moderate to severe acute pain within the adult ED. The modified version, which we call the APS-POQ-RED uses nine questions to measure three subscales (pain relief and satisfaction, affective distress and pain interference subscales). The modification of the instrument to focus on care received in the ED, as well as the removal of questions not relevant have made this a specific, validated tool for use in quality improvement and research activities within the ED.

## Supporting information

Supplementary Figure One

## Data Availability

All data produced in the present study are available upon reasonable request to the authors.

## References

1. Hughes JA, Douglas C, Jones L, Brown NJ, Nguyen A, Jarugula R, et al. Identifying patients presenting in pain to the adult emergency department: A binary classification task and description of prevalence. medRxiv. 2022:2022.05.29.22275652.

2. Cordell WH, Keene KK, Giles BK, Jones JB, Jones JH, Brizendine EJ. The high prevalence of pain in emergency medical care. The American Journal of Emergency Medicine. 2002;20(3):165–9.

3. Wong A, Potter J, Brown NJ, Chu K, Hughes J. Patient-Reported Outcomes of Pain Care Research in the Adult Emergency Department: A Scoping Review.. 2020.

4. Jao K, McD Taylor D, Taylor SE, Khan M, Chae J. Simple clinical targets associated with a high level of patient satisfaction with their pain management. Emergency Medicine Australasia. 2011;23(2):195–201.

5. Taylor D, Lee M, Johnson OG, Ding JL, Ashok A. Patient satisfaction with their pain management: The effect of provision of pain management advice. Emergency Medicine Australasia. 2016;28:8–9.

6. Gordon DB. Acute pain assessment tools: let us move beyond simple pain ratings. Current Opinion in Anesthesiology. 2015;28(5).

7. Hatherley C, Jennings N, Cross R. Time to analgesia and pain score documentation best practice standards for the Emergency Department–A literature review. Australasian Emergency Nursing Journal. 2016;19(1):26–36.

8. Hughes JA, Alexander KE, Spencer L, Yates P. Factors associated with time to first analgesic medication in the emergency department. Journal of Clinical Nursing. 2021;30(13-14):1973–89.

9. Hughes JA, Alexander KE, Spencer L, Yates P. Factors associated with the experience of patients presenting in pain to the emergency department. Journal of Clinical Nursing. 2022;31(9-10):1273–84.

10. Taylor DM, Valentine S, Majer J, Grant N. Discordance between patientLJreported and actual emergency department pain management. Emergency Medicine Australasia. 2021;33(3):517–23.

11. Flynn SB, Gordee A, Kuchibhatla M, George SZ, Eucker SA. Moving toward patientLJcentered care in the emergency department: PatientLJreported expectations, definitions of success, and importance of improvement in painLJrelated outcomes. Academic Emergency Medicine. 2021;28(11):1286–98.

12. Acquadro C, Berzon R, Dubois D, Leidy NK, Marquis P, Revicki D, et al. Incorporating the patient’s perspective into drug development and communication: an ad hoc task force report of the PatientLJReported Outcomes (PRO) Harmonization Group meeting at the Food and Drug Administration, February 16, 2001. Value in Health. 2003;6(5):522–31.

13. Meadows KA. Patient-reported outcome measures: an overview. British journal of community nursing. 2011;16(3):146–51.

14. Black N. Patient reported outcome measures could help transform healthcare. Bmj. 2013;346.

15. Frost MH, Reeve BB, Liepa AM, Stauffer JW, Hays RD, Group MFPROCM. What is sufficient evidence for the reliability and validity of patientLJreported outcome measures? Value in Health. 2007;10:S94–S105.

16. Hughes JA, Jones L, Potter J, Wong A, Brown NJ, Chu K. An initial psychometric evaluation of the APS-POQ-R in acute pain presenting to the emergency department. Australasian Emergency Care. 2021;24(4):287–95.

17. Govenment Q. Emergency departments - Performance data pre October 2020. In: Health Q, editor. Brisbane 2020.

18. Gordon DB, Polomano RC, Pellino TA, Turk DC, McCracken LM, Sherwood G, et al. Revised American Pain Society Patient Outcome Questionnaire (APS-POQ-R) for Quality Improvement of Pain Management in Hospitalized Adults: Preliminary Psychometric Evaluation. Journal of Pain. 2010;11(11):1172–86.

19. Chaw SH, Lo YL, Lee JY, Wong JW, Zakaria WAW, Ruslan SR, et al. Evaluate construct validity of the Revised American Pain Society Patient Outcome Questionnaire in gynecological postoperative patients using confirmatory factor analysis. BMC anesthesiology. 2021;21(1):1–11.

20. Fang H, Liang J, Hong Z, Sugiyama K, Nozaki T, Kobayashi S, et al. Psychometric evaluation of the Chinese version of the revised American Pain Society Patient Outcome Questionnaire concerning pain management in Chinese orthopedic patients. PLoS One. 2017;12(5):e0178268.

21. Schultz H, Skræp U, Larsen TS, Rekvad LE, Littau-Larsen J, Schmidt SF, et al. Psychometric evaluation of the Danish version of a modified Revised American Pain Society Patient Outcome Questionnaire (APS-POQ-RD) for patients hospitalized with acute abdominal pain. 2019;19(1):117–30.

22. Erden S, KaradaLJ M, Güler Demir S, Atasayar S, Opak Yücel B, Kalkan N, et al. Cross-cultural adaptation, validity, and reliability of the Turkish version of revised American Pain Society patient outcome questionnaire for surgical patients. 2018.

23. Botti M, Khaw D, Jørgensen EB, Rasmussen B, Hunter S, Redley B. Cross-cultural examination of the structure of the revised american pain society patient outcome questionnaire (APS-POQ-R). The Journal of Pain. 2015;16(8):727–40.

24. Sepahvand M, Gholami M, Hosseinabadi R, Beiranvand A. The Use of a Nurse-Initiated Pain Protocol in the Emergency Department for Patients with Musculoskeletal Injury: A Pre-Post Intervention Study. Pain Management Nursing. 2019.

25. Taylor DM, Fatovich DM, Finucci DP, Furyk J, Jin Sw, Keijzers G, et al. Best practice pain management in the emergency department: A cluster randomised, controlled, intervention trial. Emergency Medicine Australasia. 2015;27(6):549–57.

26. Schultz H, Abrahamsen L, Rekvad LE, Skræp U, Larsen TS, Möller S, et al. Patient-controlled oral analgesia at acute abdominal pain: A before-and-after intervention study of pain management during hospital stay. Journal of Applied Nursing research. 2019;46:43–9.

27. Kyriazos TA. Applied psychometrics: sample size and sample power considerations in factor analysis (EFA, CFA) and SEM in general. Psychology. 2018;9(08):2207.

28. Xia Y, Yang Y. RMSEA, CFI, and TLI in structural equation modeling with ordered categorical data: The story they tell depends on the estimation methods. Behavior research methods. 2019;51(1):409–28.

29. Hu L-T, Bentler PM. Evaluating model fit. 1995.

30. Fabrigar LR, Wegener DT, MacCallum RC, Strahan EJ. Evaluating the use of exploratory factor analysis in psychological research. Psychological methods. 1999;4(3):272.

31. Hair J, Black W, Babin B, Anderson R. Confirmatory factor analysis. Multivariate Data Analysis, 7th ed; Pearson Education, Inc: Upper Saddle River, NJ, USA. 2010:600–38.

32. Hair JF, Black WC, Babin BJ, Anderson RE. Multivariate data analysis: Pearson new international edition. Essex: Pearson Education Limited. 2014.

33. Malhotra NK, Dash S. An applied orientation. Marketing Research. 2010;2.

34. Schmitt TA. Current methodological considerations in exploratory and confirmatory factor analysis. Journal of Psychoeducational assessment. 2011;29(4):304–21.

35. Kapoor S, White J, Thorn BE, Block P. Patients presenting to the emergency department with acute pain: the significant role of pain catastrophizing and state anxiety. Pain medicine. 2016;17(6):1069–78.

36. Robyn McCahill, Samantha Keogh, Hughes JA. Adult pain and anticipatory anxiety assessment in the emergency department: An integrative literature review. Journal of Clinical Nursing. 2022.

37. Arikan A, Esenay FI. Active and Passive Distraction Interventions in a Pediatric Emergency Department to Reduce the Pain and Anxiety During Venous Blood Sampling: A Randomized Clinical Trial. Journal of Emergency Nursing. 2020;46(6):779–90.

38. Chen YJ, Cheng SF, Lee PC, Lai CH, Hou IC, Chen CW. Distraction using virtual reality for children during intravenous injections in an emergency department: A randomised trial. Journal of Clinical Nursing (John Wiley & Sons, Inc). 2020;29(3/4):503–10.

39. Koleck TA, Dreisbach C, Bourne PE, Bakken S. Natural language processing of symptoms documented in free-text narratives of electronic health records: a systematic review. Journal of the American Medical Informatics Association: JAMIA. 2019;26(4):364–79.

40. Koleck TA, Tatonetti NP, Bakken S, Mitha S, Henderson MM, George M, et al. Identifying Symptom Information in Clinical Notes Using Natural Language Processing. Nursing research. 2021;70(3):173–83.

41. Lilik Lestari MP, Wanda D, Hayati H. The Effectiveness of Distraction (Cartoon-Patterned Clothes and Bubble-Blowing) on Pain and Anxiety in Preschool Children during Venipuncture in the Emergency Department. Comprehensive Child & Adolescent Nursing. 2017;40:22–8.

42. Longobardi C, Prino LE, Fabris MA, Settanni M. Soap bubbles as a distraction technique in the management of pain, anxiety, and fear in children at the paediatric emergency room: A pilot study. Child: Care, Health & Development. 2019;45(2):300–5.

43. Ferreira GE, Machado GC, Shaheed CA, Lin C-WC, Needs C, Edwards J, et al. Management of low back pain in Australian emergency departments. BMJ Quality & Safety. 2019;28(10):826–34.

44. Robinson K, Lam B. Early emergency department representations. Emergency Medicine Australasia. 2013;25(2):140–6.

