## Supplementary Figure One for "Enhancing pain care with the American Pain Society Patient Outcome Questionnaire for use in the Emergency Department (APS-POQ-RED): validating a patient-reported outcome measure"

James A Hughes RN PhD^1,2^

Sarah Hazelwood^3^

Anna-Lisa Lyrstedt^2^

Lee Jones^4^

Nathan J Brown^2^

Rajeev Jarugula^3^

Clint Douglas^5,6^

Kevin Chu^2^

1. School of Nursing, Centre for Healthcare Transformation, Queensland University of Technology, Brisbane, Australia.
2. Emergency and Trauma Centre, Royal Brisbane and Women’s Hospital, Brisbane, Australia
3. Emergency Department, The Prince Charles Hospital, Brisbane, Australia.
4. School of Public Health and Social Work, Queensland University of Technology, Brisbane, Australia.
5. Faculty of Health, Queensland University of Technology, Brisbane, Australia
6. Metro North Hospital and Health Service, Brisbane, Australia.

Disclosures

This project was funded through The Emergency Medicine Foundation (Australia) grant number EMLE-166R34-2020. The authors report no conflicts / competing interests in the undertaking of this work.

### Supplementary Material

Supplementary Figure One: *Structure and factor loading of the previously described structure of the APS-POQ-R modified for the ED.*

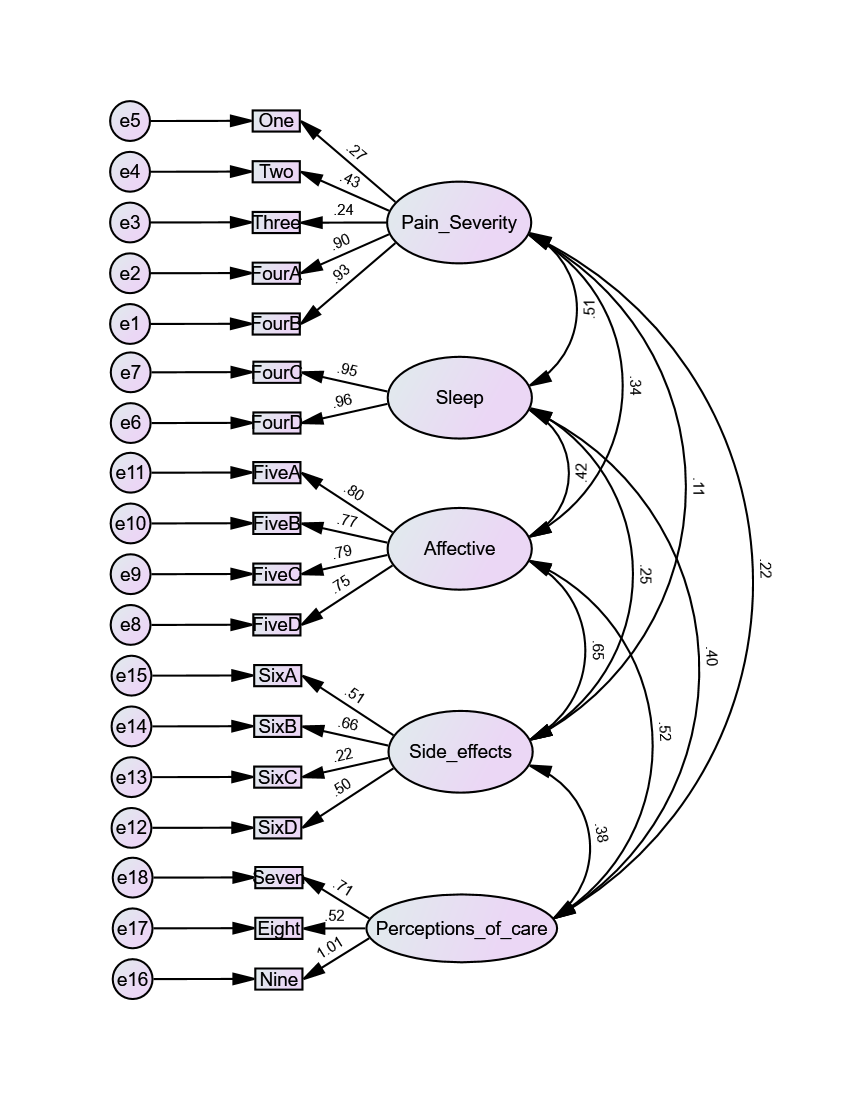

| *Supplementary Table 1*: Pattern Matrix for Exploratory Factor Analysis for objective two | | | | |
| --- | --- | --- | --- | --- |
| Number | Question | Factor | | |
|  |  | 1 | 2 | 3 |
| 1 | On this scale please indicate the least pain that you had in the emergency department? | 0.805 |  |  |
| 2 | On this scale please indicate the worst pain you had in the emergency department? |  |  |  |
| 3 | How often were you in severe in the emergency department? Please select the best estimate of the percentage of time you experience severe pain | 0.774 |  |  |
| 4a | Select one number below that best describes how much pain interfered or prevented you from Doing activities in bed such as turning, sitting up, repositioning |  |  | 0.953 |
| 4b | Select one number below that best describes how much pain interfered or prevented you from Doing activities out of bed such as walking, sitting in a chair, standing at a sink |  |  | 0.883 |
| 5a | Pain can affect our mood and emotions. On this scale, please select the one number that best shows how much the pain caused you to feel: Depressed |  |  |  |
| 5b | Pain can affect our mood and emotions. On this scale, please select the one number that best shows how much the pain caused you to feel: Frightened |  | 0.757 |  |
| 5c | Pain can affect our mood and emotions. On this scale, please select the one number that best shows how much the pain caused you to feel: Helpless |  | 0.616 |  |
| 5d | Pain can affect our mood and emotions. On this scale, please select the one number that best shows how much the pain caused you to feel: Anxious |  | 0.745 |  |
| 6a | Have you had any of the following side effects? Please select "0" if no; if yes, please circle the number that best shows the severity of each. Nausea |  |  |  |
| 6b | Have you had any of the following side effects? Please select "0" if no; if yes, please circle the number that best shows the severity of each. Drowsiness |  |  |  |
| 6c | Have you had any of the following side effects? Please select "0" if no; if yes, please circle the number that best shows the severity of each. Itching |  |  |  |
| 6d | Have you had any of the following side effects? Please select "0" if no; if yes, please circle the number that best shows the severity of each. Dizziness |  |  |  |
| 7 | In the emergency department, how much relief of your pain did you receive? Please circle the one percentage that best shows how much relief you have received from all of your pain treatments combined (medicine and non-medicine treatment) | 0.968 |  |  |
| 8 | Were you allowed to participate in decisions about your pain treatment as much as you wanted to? |  |  |  |
| 9 | Select one number that best shows how satisfied you are with the results of your pain treatment in the emergency department. | 0.805 |  |  |

|  | Pain relief and satisfaction subscale |
| --- | --- |
|  | Affective distress subscale |
|  | Pain interference Subscale |

Supplementary Table 2: *APS-POQ-RED*

**The following questions are about the pain you experienced during your stay in the emergency department**

On this scale, please tick the least pain you had in the emergency department:

From “0” - no pain to “10” - worst possible pain

| ⚪ 0 | ⚪ 1 | ⚪ 2 | ⚪ 3 | ⚪ 4 | ⚪ 5 | ⚪ 6 | ⚪ 7 | ⚪ 8 | ⚪ 9 | ⚪ 10 |
| --- | --- | --- | --- | --- | --- | --- | --- | --- | --- | --- |

How often were you in severe pain in the emergency department?

Please tick your best estimate of the percentage of time you experienced severe pain

| ⚪ 0% | ⚪ 10% | ⚪ 20% | ⚪ 30% | ⚪ 40% | ⚪ 50% | ⚪ 60% | ⚪ 70% | ⚪ 80% | ⚪ 90% | ⚪ 100% |
| --- | --- | --- | --- | --- | --- | --- | --- | --- | --- | --- |

**Tick the one number below that best describes how much this episode of has pain interfered or prevented you from different activities: (From “0” – did not interfere to “10” – completely interfered)**

|  | 0 | 1 | 2 | 3 | 4 | 5 | 6 | 7 | 8 | 9 | 10 |
| --- | --- | --- | --- | --- | --- | --- | --- | --- | --- | --- | --- |
| Doing activities in bed such as turning, sitting up, repositioning | ⚪ | ⚪ | ⚪ | ⚪ | ⚪ | ⚪ | ⚪ | ⚪ | ⚪ | ⚪ | ⚪ |
| Doing activities out of bed such as walking, sitting in chair, standing at sink | ⚪ | ⚪ | ⚪ | ⚪ | ⚪ | ⚪ | ⚪ | ⚪ | ⚪ | ⚪ | ⚪ |

**Pain can affect our mood and emotions. On this scale, please tick the number that best describes how this episode of pain caused you to feel different emotions: (From “0” – not at all to “10” – extremely)**

|  | 0 | 1 | 2 | 3 | 4 | 5 | 6 | 7 | 8 | 9 | 10 |
| --- | --- | --- | --- | --- | --- | --- | --- | --- | --- | --- | --- |
| Anxious | ⚪ | ⚪ | ⚪ | ⚪ | ⚪ | ⚪ | ⚪ | ⚪ | ⚪ | ⚪ | ⚪ |
| Frightened | ⚪ | ⚪ | ⚪ | ⚪ | ⚪ | ⚪ | ⚪ | ⚪ | ⚪ | ⚪ | ⚪ |
| Helpless | ⚪ | ⚪ | ⚪ | ⚪ | ⚪ | ⚪ | ⚪ | ⚪ | ⚪ | ⚪ | ⚪ |

In the emergency department, how much pain relief did you receive?

Please tick the one percentage that best shows how much relief you have received for all of your pain treatments combined (medicine and non-medicine treatment)

| ⚪0% | ⚪ 10% | ⚪ 20% | ⚪ 30% | ⚪ 40% | ⚪ 50% | ⚪ 60% | ⚪ 70% | ⚪ 80% | ⚪ 90% | ⚪ 100% |
| --- | --- | --- | --- | --- | --- | --- | --- | --- | --- | --- |

Tick the number that best shows how satisfied you are with the results of your pain treatment in the emergency department

From “0” – extremely dissatisfied to “10” – extremely satisfied

| ⚪ 0 | ⚪ 1 | ⚪ 2 | ⚪ 3 | ⚪ 4 | ⚪ 5 | ⚪ 6 | ⚪ 7 | ⚪ 8 | ⚪ 9 | ⚪ 10 |
| --- | --- | --- | --- | --- | --- | --- | --- | --- | --- | --- |

Adapted from: Hughes, J. A., Jones, L., Potter, J., Wong, A., Brown, N. J., & Chu, K. (2021). An initial psychometric evaluation of the APS-POQ-R in acute pain presenting to the emergency department. *Australasian Emergency Care*, *24*(4), 287-295.
